# Impact of the COVID-19 Pandemic on International Business Travel and Associated Health Issues: A Survey of Japanese Public Companies

**DOI:** 10.1101/2023.07.28.23293302

**Authors:** Yayoi Teteuou Tsukada, Ritsuko Okamura, Masahiro Yasutake

## Abstract

**Objectives:** Compared to long-term expatriates, the health issues of short-term international business travellers are less clear. Particularly, there are no reports on the impact of the COVID-19 pandemic. We explored the changes in health challenges faced by Japanese international business travellers owing to the COVID-19 pandemic.

**Design:** Cross-sectional survey research using questionnaires

**Setting:** We surveyed 3,845 listed public companies in Japan in September 2021.

**Participants:** A total of 251 companies responded (response rate: 6.5%), of which 131 (52%) had foreign travel requirements for their business.

**Primary and secondary outcome measures:** The survey included questions regarding company size, business type, necessity for foreign travel, destination and number of trips, common health issues that arise, and the importance of business travel before and after the COVID-19 pandemic.

**Results:** Among the companies, 44% replied that they could not predict the number of foreign business trips after the pandemic. However, 64% of companies responded that business travel would continue to be important in the future. Before the COVID-19 pandemic, the most important health concerns faced by business travellers were illness during travel (42%), followed by the prevention of infectious diseases and lifestyle disease management. Post-pandemic, 48% of the responses were for infectious diseases, including COVID-19, followed by 40% for travel-related diseases, and 25% for lifestyle-related diseases.

**Conclusions:** Owing to global economic and social activities, business travel will continue to be necessary in the post-COVID-19 era. Comprehensive health management including prevention of infectious diseases is desirable for business travel.

**Strengths and limitations of this study:**

- This study provides valuable insights into the health problems of short-term expatriates, an area which has been left relatively unexplored compared to its long-term counterparts.
- The potentially transformative effects of the COVID-19 pandemic on the health and well-being of expatriates are also considered.
- This study will form a foundational document for reviewing the impact of the pandemic and establishing a healthcare system designed for business travellers in preparation for future pandemics.
- The survey was exclusively conducted among Japanese firms, which restricts its scope and generalisability.
- Because the survey was conducted from managers’ perspective, it did not provide an accurate assessment of the actual health status of business travellers.

## INTRODUCTION

The globalisation of economic and social activities has led to an increase in overseas business travellers. The number of international travellers worldwide has surged from 642 million in 1980 to approximately 3.4 billion in 2019 [1]. In Japan, the number has increased from 127,000 in 1964 to approximately 20 million in 2019 [2]. Business and professional travel accounts for 14% of all international tourist arrivals worldwide [3] and 12.3% in Japan [2].

Although overseas business travellers face challenges, such as long-distance travel, jetlag [4–6], the psychological burden related to their work [7], differences in environments and cultures [8], and limited access to medical care, the health problems encountered during business travel are less precise than those encountered by expatriate employees [9,10]. Expatriate employees, who are assigned overseas for six months or longer, are tracked using visa applications; therefore, the actual number is known. Additionally, overseas resident officers need to perform medical checks before departing from their destination and after returning to Japan. However, because short-term business travellers are not legally required to notify the Ministry of Foreign Affairs or undergo health checks, it is not possible to determine their number or health status.

The COVID-19 pandemic has severely impacted overseas business travel, with the number of travellers dropping to 3.1 million in 2020 and 0.5 million in 2021. The COVID-19 pandemic disrupted overseas travellers, including restrictions on overseas travel, Polymerase Chain Reaction (PCR) testing, quarantine at entry and exit, differences in infection control measures from country to country, infection at the destination, and differences in the quality and systems of medical care.

As of September 2022, business travel has resumed as the infection has subsided. To perform their duties, it is necessary for business travellers to be healthy in their destination country. The unforeseen onset of illness at the destination can impose a considerable burden on the company, both financially and in terms of business operations. However, prior to the pandemic, both medical professionals and travellers were unaware of the need for health management when traveling abroad, and awareness-raising was necessary [11–13]. The rate of consultation and vaccination prior to travel for Japanese people traveling or on business trips overseas is low [11]. Therefore, Japanese companies need to review and support the health management of overseas business travellers after the COVID-19 pandemic [14–16]. Hence, it is necessary to understand trends in business travel and emerging health issues.

This study conducted a questionnaire survey of Japanese public companies to identify trends short-term business travel in the post-era and the health challenges they face.

## MATERIALS AND METHODS

### Survey subjects and definitions

This study was cross-sectional on-line and off-line questionnaire survey conducted and analysed according to the American Association for Public Opinion Research Standard Definitions and based on mail surveys of unnamed persons [17]. The target population was Electronic Disclosure for Investors’ Network (EDINET) [18]. This database is used as it is publicly available, covers all public companies, and is updated monthly. As departments with jurisdiction details were unavailable, the questionnaire was addressed to the Health Management Division of the General Affairs and Human Resources Department. It was sent September 1, 2021, and responses were accepted online (Google form) or by mail until December 31, 2021. Responses collected by mail were additionally entered into the online questionnaire after the deadline. The entered data content was verified by other research assistants. Involving the public in the design, implementation, reporting and dissemination programme of the study was considered, but it was not possible to work with targeted companies during the pandemic.

Short-term business travel was defined as traveling abroad for a short period (six months or less) for business or employment purposes.

The survey items were (1) the necessity of overseas business travel in the company’s operations; (2) company size (number of employees); (3) industry (mining and quarrying, gravel extraction, construction, manufacturing, electricity, gas, heat supply, and waterworks, information and communication, transportation and postal services, wholesale and retail trade, finance and insurance, real estate and rental services, academic research, professional and technical services, agriculture and forestry, fishing, accommodation and food services, lifestyle-related services and entertainment, education and learning support, healthcare and welfare, comprehensive service businesses, unclassified services, public services [excluding those classified elsewhere], and unclassifiable industries); (4) healthcare problems before the COVID-19 pandemic; (5) particularly important healthcare problems before the COVID-19 pandemic; (6) healthcare problems after the COVID-19 pandemic (dealing with COVID-19 infection, dealing with infectious diseases except for COVID-19 infection, dealing with time differences, psychological support, healthcare for lifestyle-related diseases); (7) particularly important healthcare problems after the COVID-19 pandemic; (8) problems encountered in healthcare after the COVID-19 pandemic; (9) expectations of industrial physicians and travel clinics in the post-COVID-19 era; (10) number of trips taken before the COVID-19 pandemic; (11) destinations before the COVID-19 pandemic (China and other Asian countries, North America, Europe, Russia, South America, Middle East, Pacific, Africa); (12) estimated number of trips after the COVID-19 pandemic; (13) estimated destinations after the COVID-19 pandemic; (14) importance (important, somewhat important, neither, not very important, not important) of business travel after the COVID-19 pandemic; (15) importance of business travel after the COVID-19 pandemic; (16) importance of business travel after the COVID-19 pandemic and reasons; (17) reasons or purposes for continuing to travel overseas for business after the COVID-19 pandemic (on-site technical support and guidance, business negotiations and sales, internal meetings, inspections and market research, events and conferences, delivery, repair, inspection and maintenance, training, and other).

### Data analysis

The sample size for this study was calculated using the following formula.

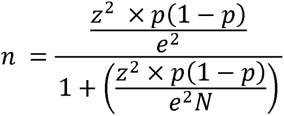

n = the required sample size (persons), N = the population size (persons), z = the confidence level (Z-score), p = the response rate (%: decimal point), e = the tolerance error (%: decimal point).

In this survey, using a tolerance error of 5%, a confidence level of 95%, and a response rate of 50%, the optimal sample size was calculated to be 254 cases.

Categorical variables were expressed as crude frequencies and percentages. The importance of health issues before and after the pandemic, number of trips, destination, and business travel were examined using a Poisson frequency test for multiple responses and Pearson’s chi-square test for single answers to determine whether there was a difference in the proportions of these variables. The two-tailed Pearson’s chi-squared test was used to test for equality of proportions. For the open-ended questionnaire, text mining analysis was performed to indicate specific word counts.

For all analyses, a two-sided p ≤ .05 was considered significant. All analyses were performed using JMP (version 14.02) statistical software (SAS Institute; Cary, NC, USA).

## RESULTS

### Background of responding companies

Among the 3,845 companies that received questionnaires, 19 returned them to an unknown address, and one refused to receive them. Of the 257 companies that responded, 5 did not indicate the company’s name, 1 declined to respond, and 251 provided valid responses (response rate: 6.5%; 2upplement 1, 2). Of the companies that responded correctly, 130 (52%) indicated that business travel was necessary for their operations. Of these, 104 (80%) were large companies with 301 employees or more. Of the 121 companies that answered that business travel was unnecessary (121 companies), 68 (56%) were small- and medium-sized companies with 300 or fewer employees. Types of industries that replied unnecessarily were the manufacturing, wholesale/retail, and service (not elsewhere classified) industries.

Eighty-five percent of companies said that overseas travel will continue to be important or somewhat important in the post-COVID-19 era. The purpose of such trips was to provide technology and guidance on-site, as well as conduct business meetings and sales (Figure 1).

**Figure 1.**
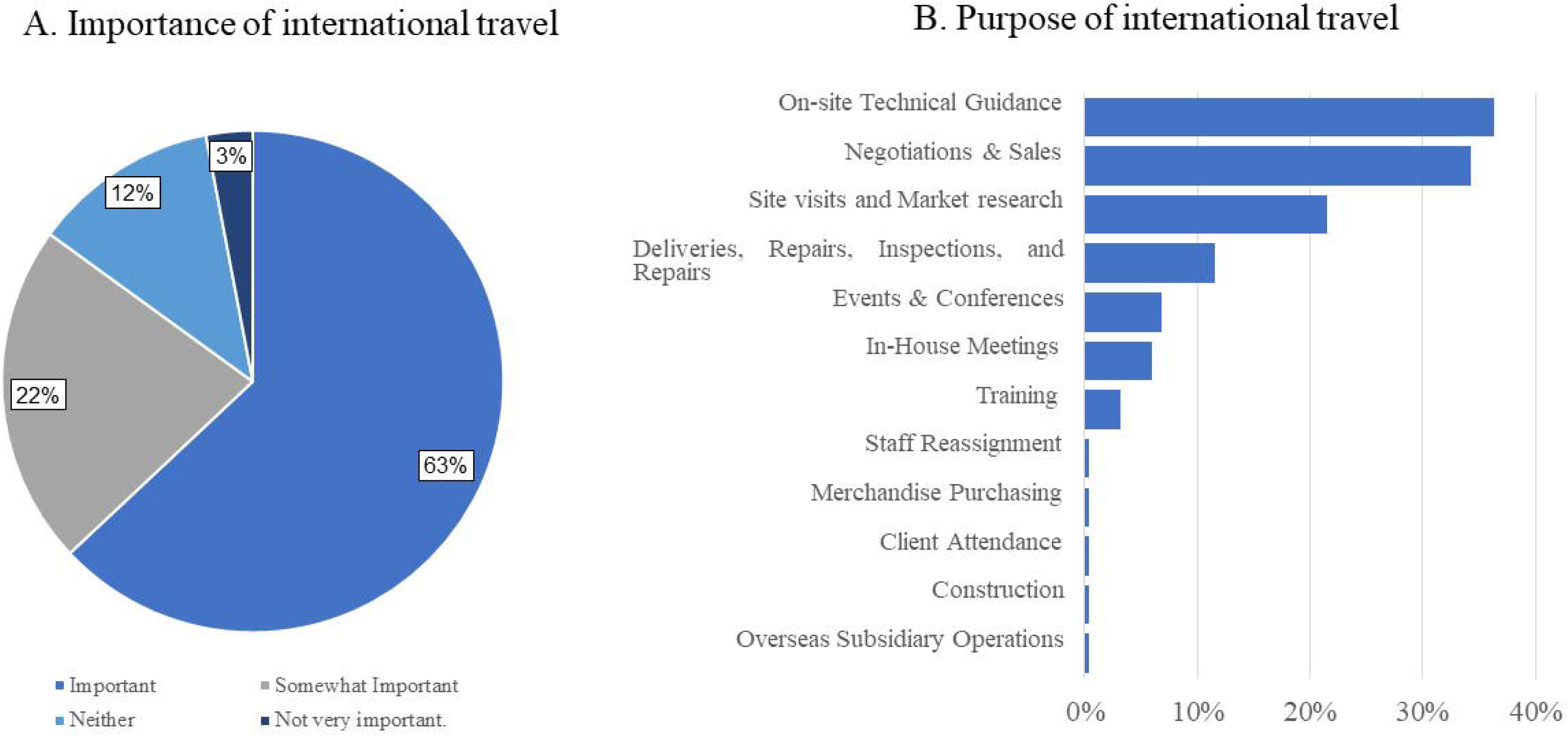
Importance and purpose of overseas travel in the post-COVID-19 era.

### Number of yearly trips and destinations before and after the COVID-19 pandemic

Before the COVID-19 pandemic, employees from 42 companies (32.3%) had travelled more than 100 times per year, and employees from 64 companies (49.2%) had travelled between 10 and 100 times. However, after the COVID-19 pandemic, 57 companies (43.8%) responded that employee travel was difficult to predict, and only 11 companies (8.5%) had more than 100 cases of employees traveling for business up until September 2021. The results indicated that the larger the company, the greater the number of trips before the COVID-19 pandemic (Pearson’s p = .0014). However, there was no difference in the estimated number of trips according to company size after the COVID-19 pandemic. Regarding the share of destinations before and after the COVID-19 pandemic, China and Asia accounted for the largest share at 40%, followed by North America, Europe, and Russia. There were differences in travel destinations by company size and industry; however, no differences existed before and after the COVID-19 pandemic (Table 1).

**Table 1.** Annual ntunber and destinations of business trips before and after COVID-19 pandemic.

|  | Before | After |
| --- | --- | --- |
| **(A) Number of yearly business travels** |  |  |
| > 500 | 13 (10%) | 1 (0.8%) |
| 300-500 | 11 (8.5%) | 1 (0.8%) |
| 100-300 | 18 (13.8%) | 9 (6.9%) |
| 10-100 | 64 (49.2%) | 22 (16.9%) |
| 1-10 | 24 (18.5%) | 34 (26.2%) |
| None |  | 6 (4.6%) |
| Unpredictable |  | 57 (43.8%) |
| **(B) Destinations** |  |  |
| Africa | 7 (2.8%) | 7 (2.8%) |
| Europe/Russia | 63 (25.1%) | 58 (23.1%) |
| Pacific | 13 (5.2%) | 13 (5.2%) |
| China/Asia | 127 (50.6%) | 125 (19.8%) |
| Middle East | 15 (6.0%) | 14 (5.6%) |
| South America | 13 (5.2%) | 14 (5.6%) |
| North America | 77 (30.7%) | 77 (30.7%) |

### Health issues for short-term business travel

Figure 2 shows the health issues associated with short-term business travel before and after the COVID-19 pandemic. The most common was ‘medical problems encountered during the business trip’ (82.2%); followed by ‘coping with infectious diseases’ (69%); and ‘general health management, such as lifestyle-related diseases’ (25 companies, 19.7%). Numerous companies (54 companies, 42.5%) cited ‘medical problems encountered during business travel’ as the most important concern (54 companies, 42.5%).

**Figure 2.**
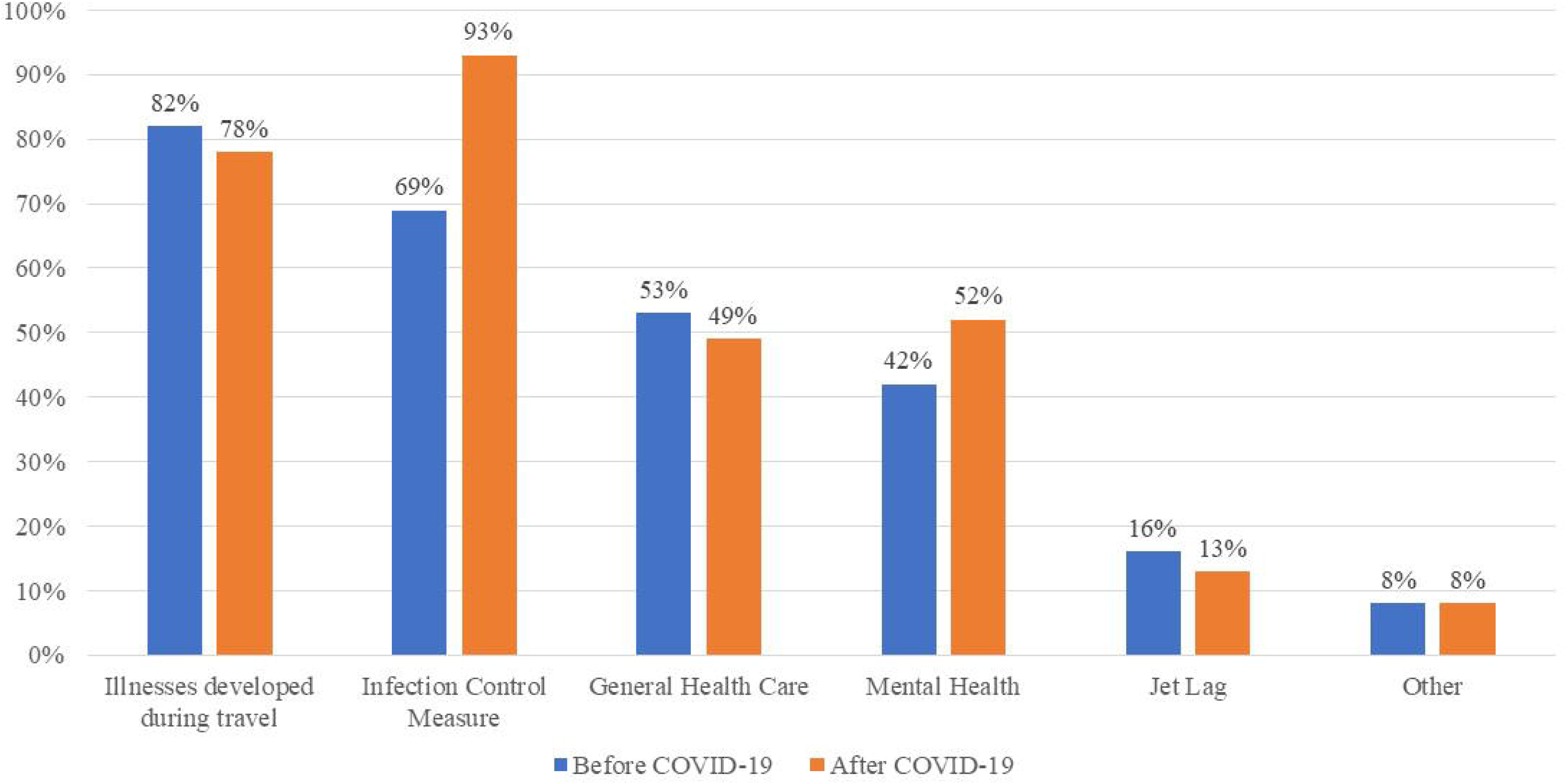
Healthcare issues of short-term Business before and after the COVID-19 pandemic (multiple answers)

Based on company size, large companies (i.e. companies with more than 300 employees) typically focus on ‘mental health management’ and ‘prevention of infectious diseases’. However, small companies (i.e. companies with 300 or fewer employees) commonly deal with ‘medical problems encountered during business travel’ and ‘general illnesses, such as lifestyle-related diseases’. ‘General health management, such as lifestyle-related diseases’ varies depending on the size of the company.

However, after the COVID-19 pandemic, the essential items were ‘prevention of COVID-19 infection’ (121 companies); ‘dealing with infectious diseases other than COVID-19’ (83 companies); ‘medical problems encountered during business travel’ (101 companies); ‘psychological support’ (67 companies); and ‘general health management, such as lifestyle-related diseases’ (63 companies), in that order (Figure 3). Numerous companies cited ‘prevention of coronavirus infection’ as the most critical factor. An analysis based on company size revealed that larger companies tended to prioritise ‘psychological support’, whereas smaller companies focused more on ‘COVID-19 infection control measures’; ‘general health management, such as lifestyle-related diseases’; ‘infection control measures against non-COVID-19 infectious diseases’; and ‘medical issues encountered during business travel’. We did not investigate differences across industries, as industries requiring overseas travel are predominantly in the manufacturing sector.

**Figure 3.**
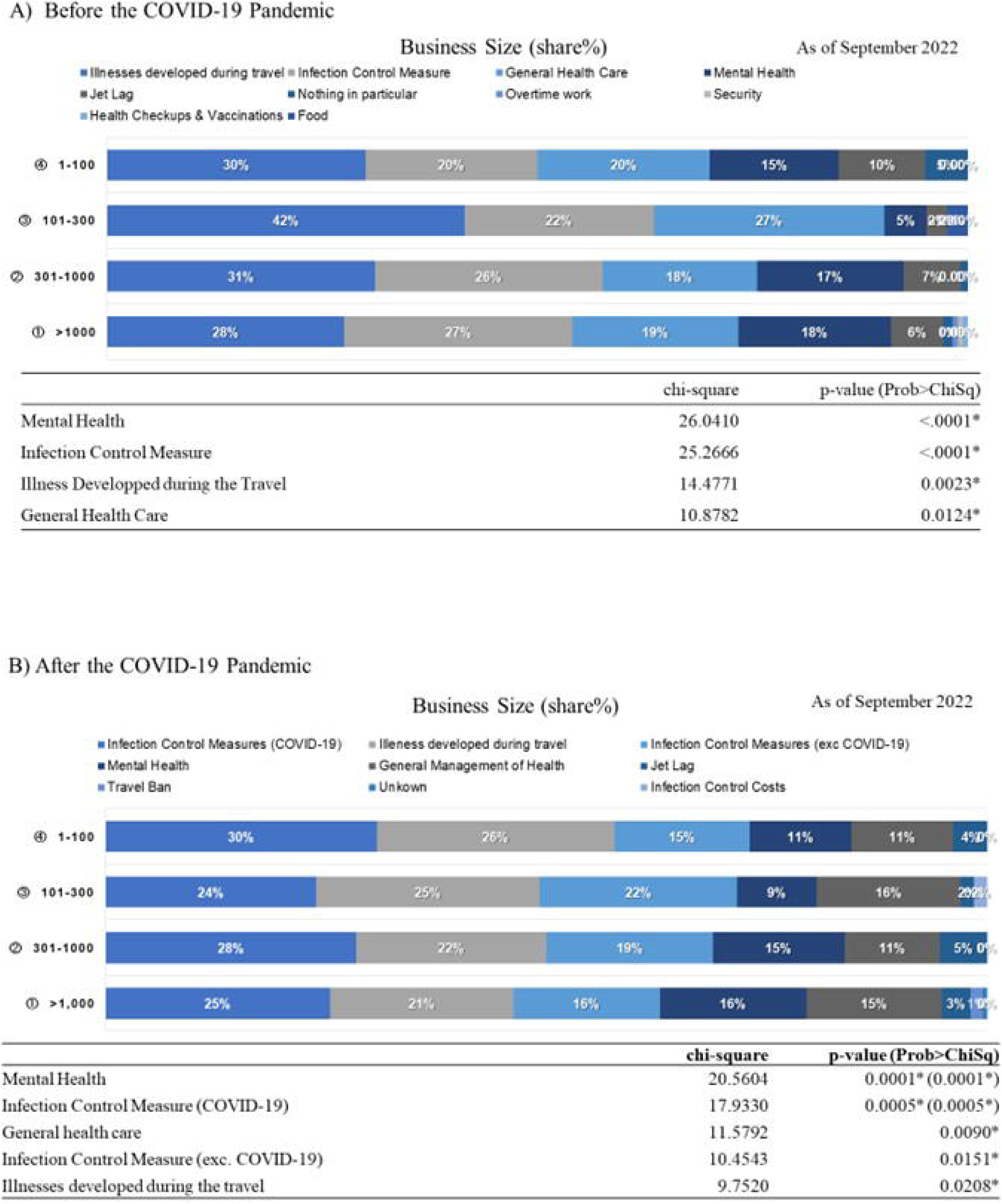
Healthcare issues ofsho1t-tenn overseas business travelers by company size.

Numerous companies mentioned vaccines during a pandemic because the survey was conducted when vaccines were difficult to obtain. Others pointed to mental health concerns, especially during quarantine, a prompt examination system, and the sharing of updated and correct information. There were comments on post-return procedures and cluster measures, the lack of regular medicine, and healthcare because of the extended quarantine. Additionally, the industrial physician/travel clinic was expected to provide guidance, advice, and judgment regarding business travel, local medical care, mental support, and medical care using information and communications technology (ICT) as well as PCR tests (Table 2).

**Table 2.**
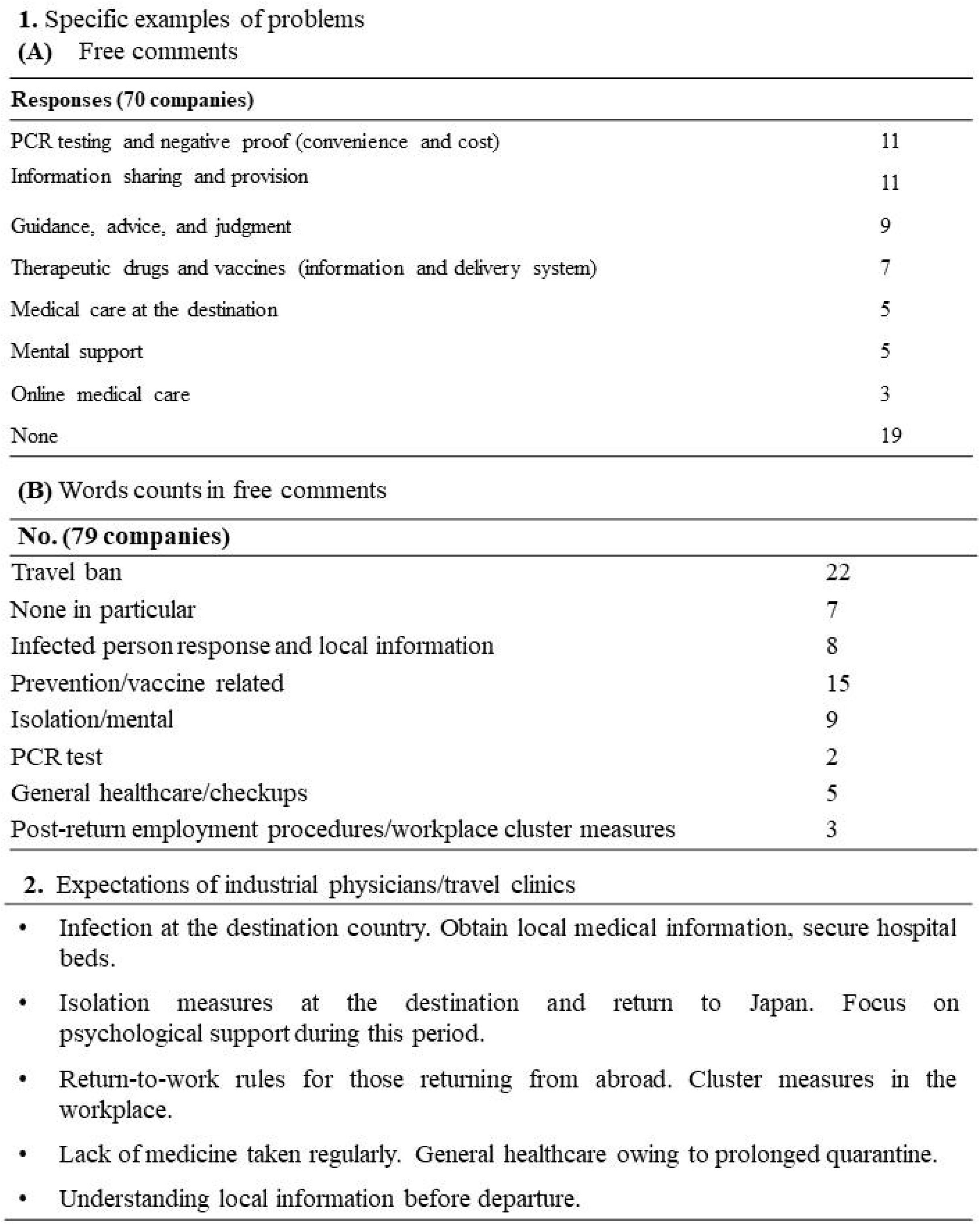
Problern cases during a COVID-19 outbreak and expectations of industrial physicians and travel clinics.

## DISCUSSION

### Trends in overseas and Japanese travel

Japan and other countries worldwide have gradually lifted quarantine restrictions relating to COVID-19 and begun accepting foreign tourists. On September 4, 2022, the Japanese government lifted all landing restrictions on individuals traveling to Japan who were previously subjected to landing denial. Visa exemption arrangements have been resumed from 0:00 am (JST) on October 11, 2022. From 0:00 am (JST) on April 29, 2023, All travellers and returnees will no longer be required to submit either a certificate of negative result of COVID-19 test conducted within 72 hours prior to departure, or a valid COVID-19 vaccination certificate of three doses or equivalent [19]. Thus, international tourism demonstrated a strong recovery through July 2022, with arrivals reaching 57% of pre-pandemic levels in the first seven months of 2022. International tourist arrivals almost tripled (+172%) in January–July 2022 compared to the same period in 2021 [1].Moreover, ICT-based alternatives such as online conferencing have become widely available. It is now easier to hold meetings and conferences without having to travel directly to a site. However, our results show that many businesses still require direct on-site operations, such as audits and guidance. Business travel decreased but remains important in the post-COVID-19 era.

### Health issues of short-term business travellers in the pre-COVID-19 pandemic period

Prior to the COVID-19 pandemic, only a few reports assessed the health problems of short-term international business travellers. In Japan, employers are legally obligated to conduct pre- and post-deployment medical examinations for those who have been sent overseas for six months or longer (Occupational Safety and Health Law). However, short-term overseas business travellers are not obligated to undergo health checkups related to their business trips, and the management of pre-trip health checkups is delegated to companies. Pre-travel health education, vaccination, and other measures for communicable diseases are optional. The number of short-term overseas travellers was greater than the number of long-term overseas travellers. However, it is difficult to identify and manage such health problems. Therefore, insufficient action has been taken to address this issue. According to a report in another country, business travellers are predominantly men, generally older, and seek pre-travel consultations largely on the advice of their employer [19,20]. The most important health issue for overseas business travellers during the pre-pandemic period was the medical problems encountered during business travel. In Japan, work accident insurance and public insurance cover some cases, but only for limited cases. Temporarily, companies must bear the full costs of both medical care and transportation. The illness of business travellers is a considerable burden for companies. Protecting business travellers’ health involves corporate responsibility and risk management.

Infectious disease control, which was previously a secondary issue, has become the biggest challenge for companies during the COVID-19 pandemic. Previously, the importance of vaccination and pre-departure consultation has been discussed and various initiatives in relation to preventable infectious diseases for individual business travellers have been reported [21–31]. Compared with other travellers, business travellers receive pre-travel healthcare closer to their travel date, and they commonly refuse vaccinations such as influenza, meningococcal, and hepatitis B vaccines [19]. In Japan, last-minute rushes in medical examinations are a problem, similar to other countries. After the pandemic, further careful measures for infection control are required, such as vaccination and its certifications. The results indicate that companies were required to take new measures since the pandemic, such as monitoring the infection situation in the destination country, implementing a return-to-work policy for those returning from abroad, and implementing cluster measures in the workplace. The Ministry of Health, Labor, and Welfare of Japan has formulated the ‘Guidelines for New Influenza Preparedness in Enterprises and Workplaces’. In preparation for future pandemics, companies are encouraged to formulate countermeasures in advance according to the pandemic phase [32]. In addition, before the COVID-19 pandemic, many respondents, primarily large companies, emphasised the importance of general healthcare, such as hypertension and diabetes. The relationship between business travel and lifestyle-related diseases has not yet been established. Previous studies reported that non-travellers were more likely to report poor/fair health compared with light travellers (1–6 nights per month), and with increasing travel, reaching 2.61 among extensive travellers (>20 nights per month). Compared with light travellers, the odds ratios for obesity were the highest among non-travellers and extensive travellers [33]. Another study indicated stronger associations between the sum of domestic and international travel and body mass index, body fat percentage, and visceral adipose tissue in women than in men after accounting for age, exercise, and sleep. Based on the male sample population, international travel frequency has a greater influence on adiposity than summed (mostly domestic) travel [34].

Conversely, an analysis of 12,942 unique health risk appraisal records of US employees of a multinational corporation indicated that international business travel was significantly associated with a lower body mass index and lower blood pressure. Generally, it has been noted that social jetlag, such as in shift workers, is associated with metabolic syndrome and cardiovascular events [35–37]. To date, there has been no analysis of the number of business trips, obesity, and metabolism among the Japanese population. Further studies are required to clarify the relationship between short-term business trips and lifestyle-related diseases. Given the lack of opportunities for health checkups and increased cost burden for companies in developing diseases overseas, companies must adequately manage their general health.

Finally, psychological support should be provided. Several studies have reported the importance of emotional support for international business travellers. A survey on health management measures among business travellers who had travelled overseas for less than six months revealed that approximately 30% of respondents experienced mental health problems or work-related disasters [38,39]. In addition, our study determined that psychological challenges were more common in larger firms. A correlation between company size and psychological stress has not been previously reported. Despite no differences in travel region or company size or industry for the responding companies, it is believed that the nature of the traveller’s job and weight of their responsibilities could be contributing factors. Balancing work and family life can also pose a psychological burden for business travellers [40,41], and a significant correlation between travel frequency and family–work conflict has been reported. Therefore, in recent years, the effective utilisation of human resources and employee assistance programmes has been suggested as a solution to help address this issue [42–44].

Upon comparing 2,962 overseas business travellers and 9,980 non-business travellers, overseas business travel was significantly associated with low body mass index, low blood pressure, excessive alcohol consumption, lack of sleep, and loss of confidence in keeping up with the pace of work [42]. The survey results indicate that during a pandemic, the psychological burden on business travellers is high owing to unexpected infection and isolation at the destination as well as differences in the medical system. ICT is helpful for workers abroad to share correct information and psychological support in a timely manner.

Thus, global companies are required to consider general occupational health management and industrial hygiene related to overseas business travel. The International Society of Travel Medicine offers travel clinics, their members, and a network of physicians who can manage overseas business travellers. In addition, the main curriculum for training industrial physicians in Japan mandates health education for those sent overseas. As overseas business travel has resumed, it may be necessary to review the health management of business travellers.

### Limitations

The survey had a notably low responding rate of 6.5%. Possible reasons for this could include employees working remotely due to the pandemic and uncertainty regarding where the company mail should be delivered. However, considering that the appropriate sample size is 254 cases as mentioned before, we believe that, statistically, it accurately reflects the population. Additionally, this survey was conducted from the perspective of managers; therefore, the actual health status of business travellers could not be ascertained. Subsequently, it is necessary to further investigate the actual health status of business travellers at companies with frequent overseas business trips. The current conclusions should be taken in the light of these limitations. However, these findings provide a meaningful step in understanding the health concerns of short-term expatriates and the impact of COVID-19 on this demographic.

## CONCLUSIONS

Considering the health issues of business travellers, it was revealed that accidents and illnesses during business trips, prevention of infectious diseases, general health management, and psychological support are essential, and that what is required differs depending on the size of the company. As overseas business travel has resumed, the health management of business travellers should be reviewed.

## Supplementary Materials

The questionnaire in English (translated from the original Japanese) can be downloaded.

## Author Contributions

Conceptualisation, Y.T.T. and M.Y.; methodology, Y.T.T.; validation, R.O. writing—original draft preparation, Y.T.T. All authors have read and agreed to the published version of the manuscript.

## Funding Statement

This research was funded by the Japan Society for the Promotion of Science Grant-in-Aid for Scientific Research (KAKENHI), grant number JP19K079901.

## Institutional Review Board Statement

This study was conducted in accordance with the Declaration of Helsinki and approved by the Ethics Committee of Nippon Medical School (no. 626-3-21).

## Informed Consent Statement

A document outlining the objectives of this study accompanied the questionnaire. Upon reviewing the document, the designated representative completed and submitted the questionnaire on behalf of their company. Consequently, we confirm that the participating companies provided their responses with consent.

## Data Availability Statement

The data presented in this study are available on request from the corresponding author. The data are not publicly available because all responses were provided in Japanese only.

## Acknowledgements

The authors thank Ms. Naoko Maekawa for providing administrative support.

## Conflicts of Interest

The authors declare no conflict of interest. The funder had no role in the design of the study; in the collection, analyses, or interpretation of data; in the writing of the manuscript; or in the decision to publish the results.

## Notes

### Competing Interest Statement

The authors have declared no competing interest.

### Funding Statement

This research was funded by Japan Society for the Promotion of Science (JSPS) Grant-in-Aid for Scientific Research (KAKENHI), Grant Number JP19K079901

