## Supplement file for "Impact of the COVID-19 Pandemic on International Business Travel and Associated Health Issues: A Survey of Japanese Public Companies"

**Survey of Health Care Issues for Short-Term Overseas Business Travelers in Post-Corona**

**Q1: Does your business require overseas travel services?**

- Necessary	　　　　　　Not necessary

**If you selected "necessary" in Q1, please answer the following questions (Q2-Q13).**

**If you chose "not required," please answer questions A-C.**

**Q2.** **What were the health care needs of short-term international travelers in before the spread of coronavirus infection** (multiple responses allowed)?

□ Diseases that may occur during the travel period

□ Prevention of infectious diseases

□ Time difference

□ Mental health management

□ General health care, including lifestyle-related diseases

□Other (　　　　　　　　　　　　　　　　　　　　　　　　　　　　　　　　　　　　　　　　　)

**Q3. Which of these were particularly important to you? Please select one.**

□ Diseases that may occur during the travel period

□ Prevention of infectious diseases

□ Time difference

□ Mental health management

□ General health care, including lifestyle-related diseases

□Other (　　　　　　　　　　　　　　　　　　　　　　　　　　　　　　　　　　　　　　　　　)

**Q4: What are the future needs in terms of health care for short-term international travelers after the spread of coronavirus infection? (Multiple answers allowed)**

□ Coronavirus infection control

□ Response to infectious diseases other than coronavirus

□ Diseases that may occur during the travel period

□ Time difference

□ Mental support

□ General health care, including lifestyle-related diseases

□ Other ( 　　　　　　　　　　　　　　　　　　　　　　　　　　　　　　　　　　　　　　　　)

**Q5. which of these have become particularly important to you? Please choose one.**

□ Coronavirus infection control

□ Response to infectious diseases other than coronavirus

□ Diseases that may occur during the travel period

□ Time difference

□ Mental support

□ General health care, including lifestyle-related diseases

□ Other ( 　　　　　　　　　　　　　　　　　　　　　　　　　　　　　　　　　　　　　　　)

**Q6. Please tell us about any particular problems that have arisen in the health care of short-term international travelers as a result of the spread of coronavirus infections.**

**Q7. In the future, what do you expect to see in the health care of short-term international travelers by industrial physicians and travel clinics?**

**Q8: Please provide an approximate number of your company's annual business travel abroad prior to the spread of coronavirus infection.**

□1-10 cases □ 10-100 cases □ 100-300 cases □ 300-500 cases □ 500 or more cases

**Q9. What are your main travel destinations?** **(Multiple answers allowed )**

□ China and Asia	□ North America

□ Europe and Russia	□ South America

□ Middle East	□ Pacific

□ Africa Region

**Q10. Please let us know the annual number of business travel abroad in the post-Corona period (forecast)**.

□ Difficult to predict □ Will disappear

□ 1-10 cases □ 10-100 cases □ 100-300 cases □ 300-500 cases □ 500 or more cases

**Q11: What are the main destinations of the post-Corona period? (Multiple answers allowed)**

□ China and Asia	□ North America

□ Europe and Russia	□ South America

□ Middle East	□ Pacific

□ Africa Region

**If you answered in Q10 that you will resume business overseas travel in the post-Corona period, what is your reason or purpose for continuing your business overseas travel? (Multiple answers allowed)**

**□** On-site technical support and guidance	□ Commercial talks and sales

□ Internal meetings	□ Market research

□ Events and conferences	□ Delivery, repair, inspection, repair

□ Training

　Other ( )

**Q13: What is the importance of short-term overseas travel for overseas business in the post-Corona period?**

**□** Will continue to be important □ Somewhat important □ Neither

□ Not very important □ Not important

Reason

**A: How many employees does your company have?**

- 1-100 persons □ 101-300 persons □ 301-1000 persons □ 1,001 persons or more

**B: What is your company's type of business?**

- Mining, Quarrying, Gravel extraction
- Construction
- Manufacturing
- Electricity, gas, heat supply, and water supply
- Information and Communication
- Transportation, postal service
- Wholesale and retail
- Finance, Insurance
- Real estate, goods rental
- Academic research, professional and technical services
- Agriculture, Forestry
- Fishing
- Lodging, Restaurants
- Lifestyle-related services, recreation
- Education, Learning Support
- Medical, Welfare
- Complex service business
- Services (not elsewhere classified)
- Public affairs (excluding those classified elsewhere)
- Unclassifiable Industries

**C: Please provide any other comments or concerns about health care for short-term business travelers.**

**Company Name**:

**Name of person in charge**:

**Thank you very much for your cooperation.**
