## Supplementary figures and images for "Impact of the COVID-19 Pandemic on International Business Travel and Associated Health Issues: A Survey of Japanese Public Companies"

### Supplement1

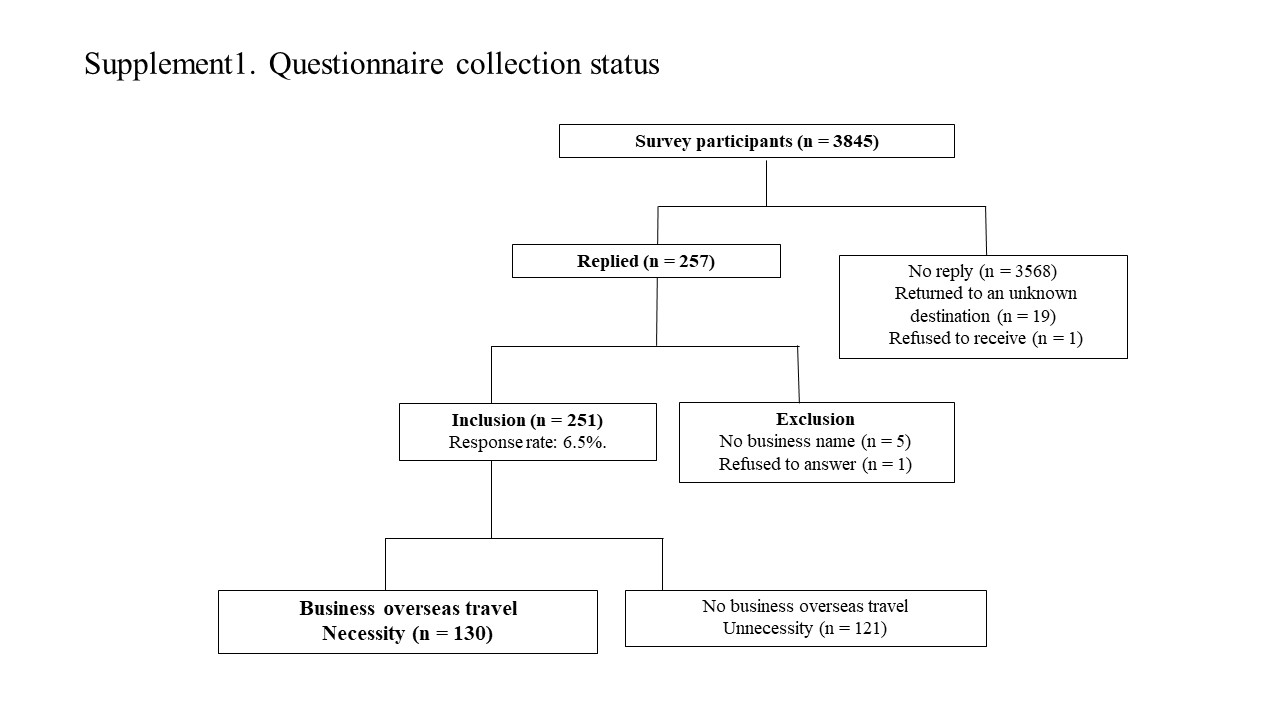

### Suppliment2

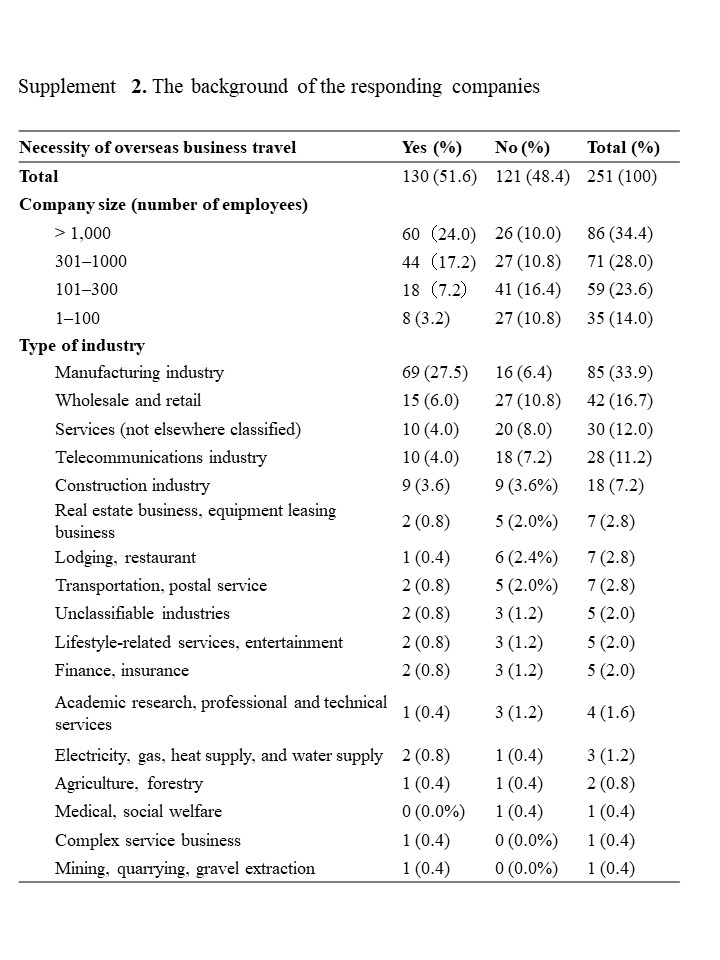
